# Environmental circulation of adenovirus 40/41 and SARS-CoV-2 in the context of the emergence of acute hepatitis of unknown origin

**DOI:** 10.1101/2022.06.08.22276091

**Authors:** Elke Wollants, Els Keyaerts, Lize Cuypers, Mandy Bloemen, Marijn Thijssen, Sien Ombelet, Joren Raymenants, Kurt Beuselinck, Lies Laenen, Lore Budts, Bram Pussig, Katrien Lagrou, Marc Van Ranst, Emmanuel André

## Abstract

The recent surge of hepatitis of unknown origin in children is hypothesized to be caused by adenovirus 41 and/or SARS-CoV-2 infections. A relatively high proportion of patients testing positive for these viruses concomitantly with the development of acute hepatitis supports this hypothesis. To formally incriminate these viral infections as causative agents of hepatitis, both a plausible physiopathological pathway and supporting epidemiological dynamics in the community need demonstration. In this study, we measured the level of circulation of adenovirus 40/41 and SARS-CoV-2 in the general population of the city of Leuven in Belgium using wastewater monitoring between December 2020 and May 2022 and indoor air sampling in day care centers between November 2021 and May 2022. We also retrospectively analyzed medical records of 12.672 children attending a tertiary hospital draining the same region between January 2019 and April 2022. Our results demonstrate a recent but modest increase in hepatitis of unknown origin concomitant with a surge of circulating adenovirus 41 and SARS-CoV-2 in the general population, including in children under 5.

## The study

To date, 650 probable cases of acute hepatitis of unknown etiology in children have been reported to the WHO. Three out of four (75.4%) of these cases are in children <5 years old. The majority of reported cases (n=374; 58%) are from the WHO European Region. In total, 181 cases were tested for adenovirus (AdV), of which 110 (60.8%) tested positive and 188 cases were tested for SARS-CoV-2, of which 23 (12.2%) tested positive. To date, Belgium reported 14 probable cases [1].

The etiology of these severe acute hepatitis remains unknown and under investigation. It is plausible that adenovirus infection plays a role in the physiopathology of this disease, potentially in synergy with SARS-CoV-2.

COVID-19 associated hepatitis has been reported as an uncommon clinical entity [2], [3]. The continued high circulation of SARS-CoV-2 among children may reveal rare and serious consequences of this disease, even if a COVID-19 infection is generally considered to be mild in children [4].

Adenovirus 40/41 is one of the leading causes of childhood diarrheal disease worldwide [5]. Its contribution to the current outbreak should be based on both a plausible physiopathological pathway and supporting epidemiological dynamics rather than uncontextualized documentation of infections in patients with a hepatitis of unknown origin.

We hypothesize that, if adenovirus and SARS-CoV-2 were physiopathological contributors, we should be able to demonstrate a link between the intensity of (co-)circulation in a population and the number of cases of hepatitis of unknown origin emerging in this same population. This study describes the retrospectively assessed dynamics of adenovirus 40/41 and SARS-CoV-2 community transmission at the level of Leuven and sub-municipalities (115.000 inhabitants) in Belgium. In parallel, we systematically identified cases of acute hepatitis in children under 16 years old at UZ Leuven University Hospitals, whose catchment area covers the city and a network of 31 regional hospitals, including a referral center for liver transplantation in children [6], [7].

### Circulation of adenovirus 40/41 and SARS-CoV-2 in the general population in wastewater

We retrospectively assessed the level of circulation of adenovirus and SARS-CoV-2 by performing AdV 40/41 and SARS-CoV-2 qPCR on 63 sequential wastewater samples collected between 06/12/2020 and 16/05/2022. These samples were taken at a sampling site allowing the surveillance of SARS-CoV-2 transmission in the entire city [8]. As presented in Figure 1, the circulation of SARS-CoV-2 peaked in April 2021 (Alpha wave) and December 2021 (Delta wave), and thereafter remained at a high level throughout the Omicron BA.1 (January 2022) and BA.2 (March 2022) waves of infection. A considerable surge of AdV 40/41 was observed between July and September 2021, and the circulation remained at high level thereafter. AdV subtyping was attempted on all positive sewage samples using Sanger sequencing. Among the samples for which subtyping was possible (57%), AdV 41 was detected in 65% of the samples, and was only observed after the July 2021 surge. The positivity rate and mean Ct value of adenovirus and SARS-CoV-2 in wastewater are shown in table1.

**Figure 1:**
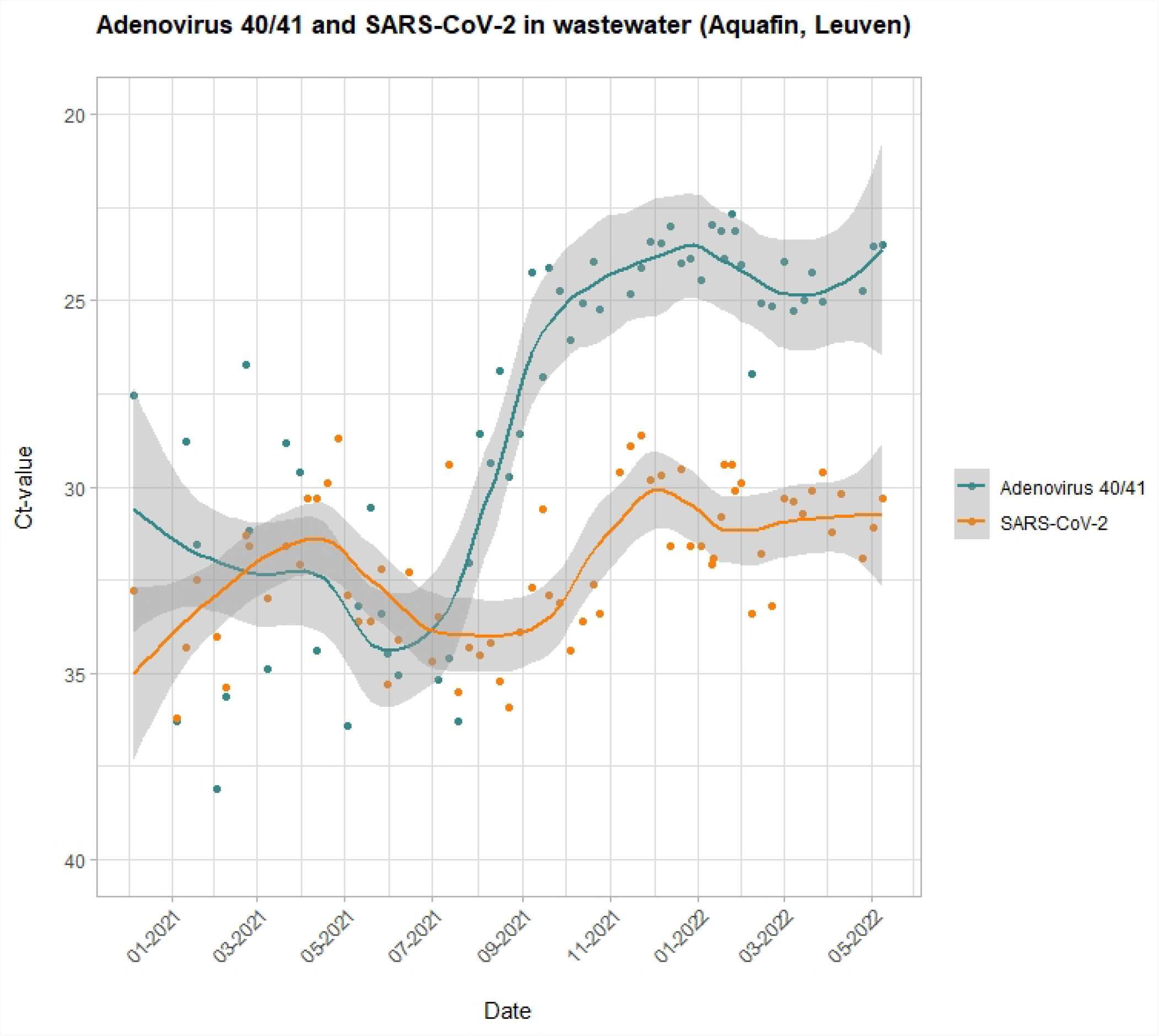
Evolution of AdV 40/41 and SARS-CoV-2 circulation in the general population between December 2020 and May 2022. The intensity of the signal is captured by the Ct value of targeted PCR assays performed on wastewater.

**Table 1:**
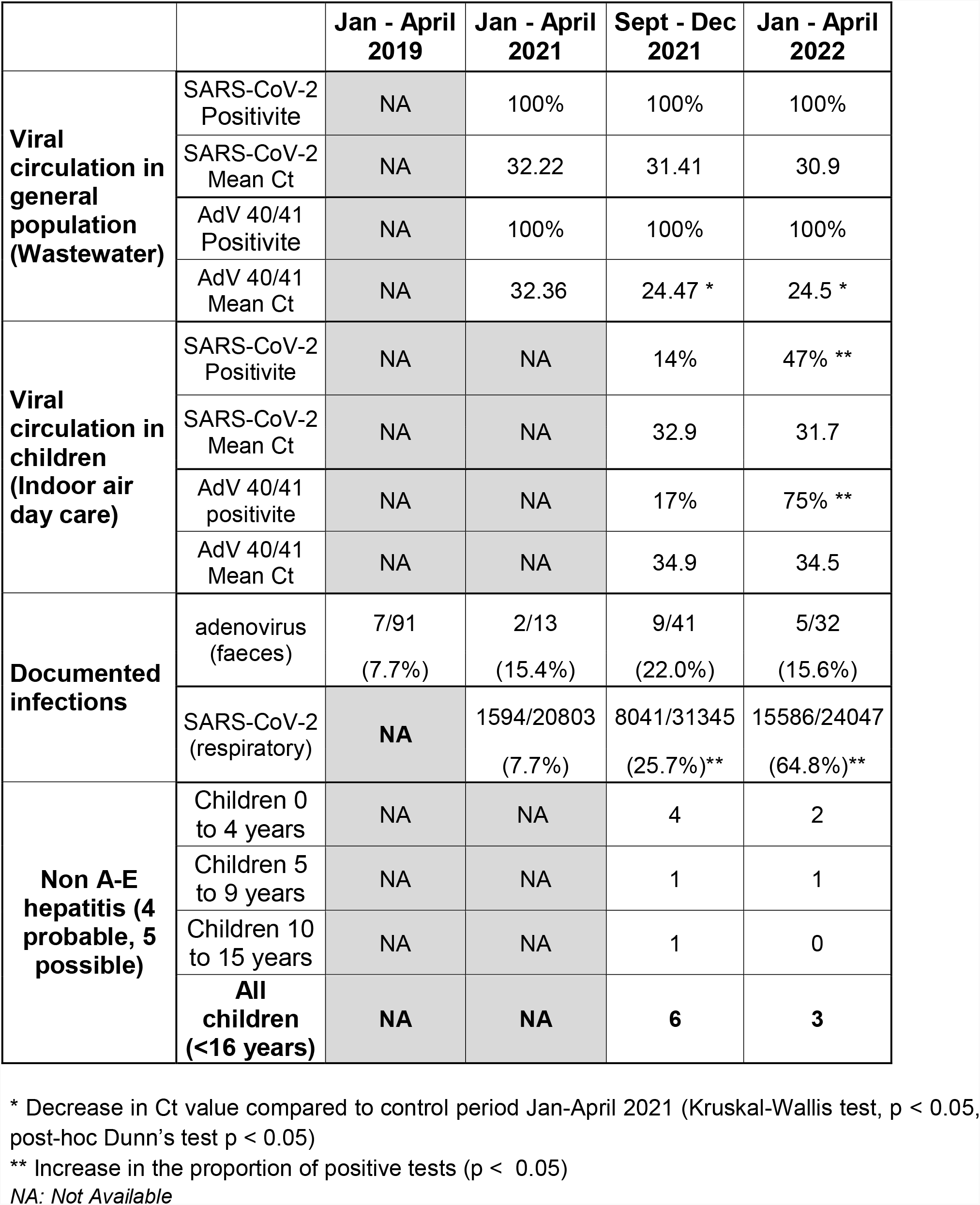
Overview of adenovirus 40/41 and SARS-CoV-2 circulation in the environment (wastewater and indoor air samples from children day care centers), laboratory-confirmed infections with both viruses in children under 16 referred to the University Hospital Leuven. By verifying medical records of children with elevated liver enzymes, 4 probable and 5 possible non A-E hepatitis cases of unknown origin have been identified since October 2021

**Table 2:** Characteristics of the 9 children (out of 12.672 screened) which presented with increased liver enzymes where acute viral hepatitis was the most likely differential diagnosis between October 2021 and April 2022. The analyses were not systematically performed at diagnosis (date of onset hepatitis = date of documented elevated liver enzymes). They were performed in addition if sufficient sample material was still available at the time of supplementary testing. In the context of hepatitis screening, the following analyses were performed: IgM and IgG antibodies (Abs) for hepatitis A, C and E virus (HAV, HCV and HEV) and antigen (Ag) and Abs for surface and core hepatitis B virus (HBV). If testing was not performed at admission and the correct specimen type was unavailable in sufficient volume, this is indicated as ‘no sample available’ and highlighted in gray. ^1^ Patient 2: immune for HAV (IgG positive and IgM negative) and HEV (IgG positive and IgM negative); and vaccinated for HBV (Ag and core Abs negative, while surface Abs positive) ^2^ Patient 7: vaccinated for HBV (Ag and core Abs negative, while surface Abs positive) * All patients recovered and were discharged from the hospital

|  | Age | Date onset hepatitis | Hepatitis screening | Adenovirus 40-41 | SARS-CoV-2 | Clinical evolution |
| --- | --- | --- | --- | --- | --- | --- |
| Patient 1 | 0-4 | October 2021 | No sample available | No sample available | No sample available | Recovery* |
| Patient 2 | 10-15 | October 2021 | Non A-E (Immune for HAV, HBV-HEV)<sup>1</sup> | Negative | Negative | Recovery* |
| Patient 3 | 0-4 | October 2021 | No sample available | Negative | Negative | Recovery* |
| Patient 4 | 0-4 | November 2021 | No sample available | Negative | Negative | Recovery* |
| Patient 5 | 5-9 | December 2021 | Non A-E | No sample available | Negative | Recovery* |
| Patient 6 | 0-4 | December 2021 | No sample available | No sample available | No sample available | Recovery* |
| Patient 7 | 0-4 | January 2022 | Non A-E (Immune for HBV)<sup>2</sup> | Negative | Negative | Recovery* |
| Patient 8 | 5-9 | January 2022 | Non A-E | Negative | Positive January 2022 | Recovery* |
| Patient 9 | 0-4 | March 2022 | No sample available | Negative | Negative | Recovery* |

### Adenovirus 40/41 and SARS-CoV-2 circulation in children under 5 in indoor air samples

To assess the level of circulation of Adenovirus 40/41 and SARS-CoV-2 in children under 5 years of age in the general population, we analyzed 158 air samples collected in children day care centers located in the city of Leuven (128 samples from crèches and 30 from kindergarten), collected between November 2021 and May 2022 [9]. These samples were systematically collected as part of a prospective study aiming to investigate co-circulation of respiratory pathogens in children day care centers. As shown in Figure 2, adenovirus 40/41 was detected for the first time in the air on December 10^th^, until then 15 air samples tested negative for adenovirus 40/41. Subsequently, both adenovirus 40/41 and SARS-CoV-2 were consistently detected in the indoor air of children day care facilities throughout the evaluated period, which is concomitant with the period during which cases of hepatitis were reported in Belgium and around the world. The positivity rate and mean Ct value of adenovirus and SARS-CoV-2 in indoor air samples are shown in table1.

**Figure 2:**
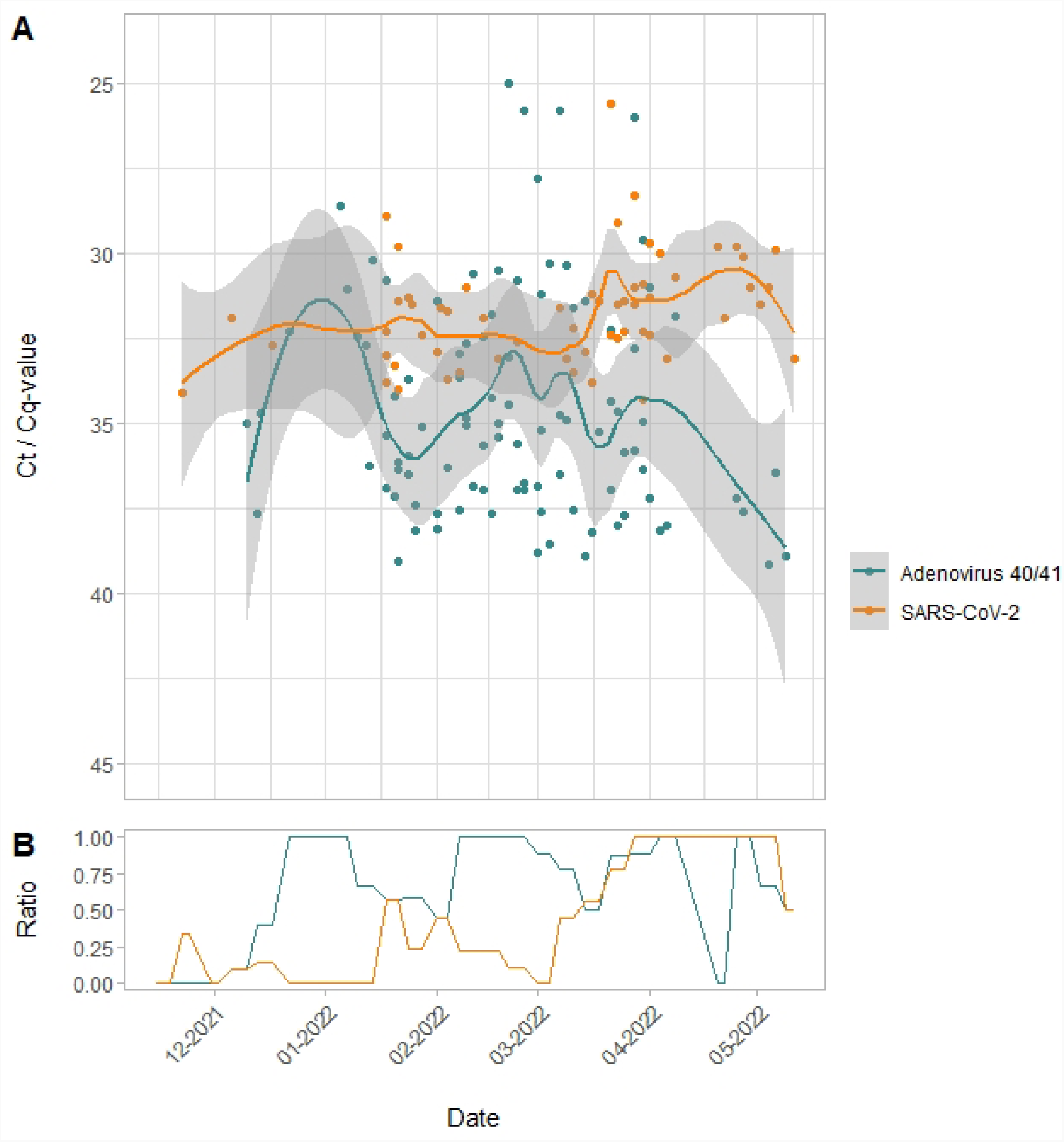
A: Evolution of Adv 40/41 and SARS-CoV-2 circulation in children day care centers between November 2021 and May 2022. The intensity of the signal is captured by the Ct / Cq value of targeted PCR assays performed on air samples. B: The ratio of positive samples to the total of tested samples is depicted in lower figure.

### Documented adenovirus 40/41 and SARS-CoV-2 infections in children under 16

We retrospectively looked for documented adenovirus and SARS-CoV-2 infections in children under the age of 16 years old, diagnosed in the Laboratory Medicine department of University Hospitals Leuven. Faeces and respiratory samples were included. An *in house* developed pan-adenovirus PCR and an AdV 40/41 rapid antigen test were used to diagnose an AdV infection in fecal material, while for SARS-CoV-2, all PCR assays performed according to the routinely implemented and accredited testing criteria were evaluated. Four timeframes were considered to cover pre- and post-COVID-19 periods, results are presented in table1.

### Active case finding investigation for hepatitis of unknown origin

We retrospectively evaluated the medical records of children <16 presenting with elevated liver enzymes (serum transaminase >500 IU/L, either AST or ALT), whose sample was referred to the Laboratory Medicine department of the University Hospitals Leuven. After evaluation nine children with possible viral hepatitis were retained. Viral testing during hospitalization for hepatitis A-E, Adenovirus 40/41 and SARS-CoV-2 was checked. A complete viral screening was not systematically performed in any of the cases. In 4 out of 9 children, we could retrieve blood samples with sufficient volume for complementary testing. All nine developed hepatitis after October 2021 and recovered without requiring liver transplantation. Hepatitis A-E could be ruled out in four based on negative serology or immune status. AdV infection was ruled-out in six (by PCR on fecal and blood samples) and SARS-CoV-2 infection was ruled out in six (by PCR on respiratory samples). One had a documented SARS-CoV-2 infection four days prior to the onset of hepatitis. COVID-19 associated hepatitis was considered the most likely diagnosis in the latter.

### Conclusion

Wastewater surveillance and indoor air monitoring in children day care centers highlight high levels of adenovirus 40/41 and SARS-CoV-2 in the general population between September 2021 and April 2022. In the same period, we identified 4 probable and 5 possible cases of non-severe non-A-E viral hepatitis of unknown origin in children seeking care in a large tertiary hospital for which transaminase blood tests were available. No such cases were identified in the earlier timeframes, which spanned both the pre-COVID-19 and post-COVID-19 periods. None of these patients had a documented adenovirus infection. SARS-CoV-2 was directly incriminated in one patient. They all had a spontaneous favorable outcome and did not require liver transplantation.

Taken together, our observations do not confirm or exclude the possibility that adenovirus or SARS-CoV-2 infections are contributors to the rise in acute hepatitis cases in children. If either or both do however contribute, only a small minority of infections are likely to lead to severe hepatitis. This is supported by the high rates of community transmission of both viruses and the low number of admissions with viral hepatitis cases of unknown origin in children between September 2021 and April 2022.

This study underlines the importance of environmental monitoring in a holistic disease surveillance approach. In this particular context, estimating the level of circulating adenovirus and SARS-CoV-2 in the general population has shown to be critical for estimating the frequency of eventual secondary hepatitis associated with these viruses. If a causality would be suggested by other studies, our results indicate that severe hepatitis cases would only emerge in the context of massive viral circulation in a very large population.

## METHODS

### Sample collection

Aquafin is a company in Flanders (Belgium) with a wastewater treatment plant (WWTP) that collects and treats municipal wastewater. The treatment plant covers an area of approximately 115000 inhabitants within 8 different municipalities. Weekly samples (500 mL) of 24-hour composite influent wastewater were collected through a time-proportional automated sampler. This sampler collects 50 mL of wastewater every 10 minutes in a large container. Before transport to the laboratory, samples were stored in a refrigerator at 4°C.

Aerosol samples were collected as described by Cuypers *et al*. 2022 [10], in congregate settings of different age groups: nursery (0-3y) and kindergarten (3-6y). Air samples from the créche were collected in the bathroom near the toilet, air samples from the kindergarten were collected in the children’s dining area. Samples were taken for 2 hours 1-3 times a week between November 2021 and April 2022 and were transferred to transport medium (UTM, Copan and InActiv Blue, Fertipro) prior to analysis.

### Detection of SARS-CoV-2 in wastewater by RT-qPCR

Wastewater samples were collected and concentrated. SARS-CoV-2 was detected as previously described in Bloemen *et al*. 2022 [8].

### Nucleic acid extraction

RNA extraction for SARS CoV-2 and AdV was performed using MagMAX™-96 Viral RNA Isolation Kit for automated extraction (Thermo Fisher Scientific, AM1836), with 200 μL sample input. Samples were spiked with a purified MS2 bacteriophage as internal control according to the manufacturer’s instructions (Thermo Fisher Scientific, A47817). Extracted RNA was eluted from magnetic beads in 50 μL of UltraPure DNase/RNase free distilled water.

Total nucleic acid (TNA) for AdV PCR is extracted from 500 µL air sample in UTM with NucliSens extraction reagents on easyMAG or eMAG (BioMérieux, Lyon, France) using the specific B protocol on the instrument after off-board lysis during 10 minutes and continuous shaking. 10 μL of a mixture of Phocine Distemper Virus (PDV) [11] and Phocine Herpesvirus-1 (PhHV-1) [12] (PDV en PhHV-1 stock kindly supplied by Groningen Medical Center, Groningen, The Netherlands) is added to the lysed sample before extraction and serve as an RNA and DNA internal control (IC) in the assay. The elution volume of TNA is 110 µL.

### Detection of SARS-CoV-2 in air samples by RT-qPCR

RT-PCR testing was performed as described by Cuypers *et al*. 2022 [10].

### Detection of adenovirus 40/41 in wastewater and air samples by RT-qPCR

To detect adenovirus in sewage water and air samples, specific primers and probes were used [13]. Five µL TaqMan™ Fast Virus 1-Step Master Mix (Applied Biosystems, CAT 4444434) was mixed with 1 µL JTVFF primer (20µM), 1 µL JTVFR primer (20µM) and 0.5 µL JTVFAP probe (10µM). Cycling conditions were 3 minutes 95°C followed by 40 cycles of 15 seconds at 96°C and 30 seconds at 60°C.

### Sanger sequencing of adenovirus 41 wastewater samples

Sanger sequencing was performed according to Bloemen *et al* [14]. Adenovirus primers, able to detect the different circulating AdV types, were used [15].

### Active case finding investigation for hepatitis of unknown origin

Four timeframes were considered to cover pre- and post-COVID-19 periods: January to April 2019 (7063 blood samples from 2938 children), January to April 2021 (7172 blood samples from 3139 children), September to December 2021 (7338 blood samples from 3188 children) and finally January to April 2022 (7535 blood samples from 3407 children).

After deduplication to individual patient level, a total of 63 children under 16 were identified with elevated liver enzymes over all periods. Their distribution was as follows: 22/2938 (0.75%) for January to April 2019, 12/3139 (0.38%) for January to April 2021, 14/3188 (0.44%) for September to December 2021 and 15/3407 (0.44%) for January to April. There was thus no significant difference between periods.

We subsequently screened medical records of these 63 children and excluded cases where viral hepatitis was not the most likely differential diagnosis.

### Statistical analysis

All statistical analyses were performed and visualized in R [16] using the ggplot2 package [17]. Changes in the number of elevated liver enzymes cases and pathogen positivity were tested with the proportions test function between the study periods. The Kruskal-Wallis test was used to test differences between groups of continuous variables, followed by a Dunn’s test for pairwise comparisons between groups. The Benjamin-Hochberg method was used for multiple testing corrections with a false discovery rate adjustment cut-off < 0.05.

## Data Availability

All data produced in the present study are available upon reasonable request to the authors

## Acknowledgements

We would like to gratefully acknowledge the support that has been provided to this study by our colleagues Hannelore De Muylder, Bastiaan Craessaerts, Gertjan Gysembergt and Camille Vanderschoot. We would like to thank Valentijn Vergote for the ICT support.

